# COVID-19 case management: The policy model in Morocco context

**DOI:** 10.1101/2020.11.07.20227603

**Authors:** Issam Bennis

## Abstract

**Background:** Based on the updated scientific evidence around SARS-CoV-2 diagnostic, health policymakers had to consider that many decisions could enhance or limit the success of the overall COVID-19 control strategy. The purpose of this study is to share alternative COVID-19 case management based on the updated international knowledge.

**Methods:** This study presents the main information about COVID-19 case management in Morocco from March to October 2020. The NVivo qualitative model content analysis was used to compare and prioritize health decisions with updated scientific evidence.

**Results:** The lack of molecular diagnostic accuracy using the interpretation of cycles quantification values, was targeted only by allowing all private laboratories to do RT qPCR. However, there is an urgent need for standardisation with accurate molecular SARS-CoV-2 thermocyclers and kits that notify systematic cycles quantification and do more tests per days to control the spread effectively. A predictive tree of the cycle’s range is presented following three steps: 1) the initial clinical definition, 2) the molecular confirmation, 3) and the diagnostic follows up results of the RT qPCR up to 28 days after the onset. At the same time, the seasonal vaccination against influenza and pneumonia could help to reduce COVID-19 deaths.

**Conclusions:** Until an available SARS-CoV-2 specific vaccine and/or curative effective treatment, updated control strategy in Morocco and similar context countries require to target population living in highly COVID-19 epidemic cities or areas by mass testing with the right interpretation of PCR values changes, associated to seasonal vaccination to foster the immunity.

## Background

Coronavirus 2019 (COVID-19) is a new infectious disease due to severe acute respiratory syndrome coronavirus type 2 (SARS-CoV-2). The internationally accepted diagnostic tool to be used to confirm SARS-CoV-2 is to quantify the viral load in each sample. Quantitative real-time Polymerase Chain Reaction (RT-qPCR) is cited as the acceptable diagnostic device used for SARS-CoV-2 diagnosis. While for some scientists, the reverse transcriptase PCR is considered as the gold standard[1]. The current diagnostic testing methods recommended by the World Health Organization for testing SARS-CoV-2 require two steps: RNA extraction from patient nasopharyngeal swabs; and RT-qPCR amplification of viral load[2].

Up to date, the molecular testing is mainly dependant on the financial budget allocated compared to the scientific advances and technics developed. Thus, the countries’ capacities to perform daily COVID-19 tests targeting many categories (affected, suspected, or undefined) become one of the most trending ways to show the robustness of their health systems. While, in some countries, primary prevention is the main prioritized strategy to control COVID-19 spread. Then, the population behaviours are spotted as the main obstacle to getting success.

For instance, Morocco, as a Middle-income country, enhanced his overall capacity of molecular testing by creating the national laboratory COVID-19 networking. Then, more than thirty public and private laboratories meet the criteria of being able to analyse SARS-CoV-2 samples in a safe environment. The increase of involved laboratories allows going from 300 maximum tests per day done in March to 9000 in May, then, 16000 in June to more than 20000 since end of July, to do not increase more than 21000 to 22000 per day during September and October 2020. Unfortunately, some collective misconduct behaviours, including less respect to the lockdown and not taking the overall preventive measures or avoiding gatherings of more than ten people earnestly, are pointed as the source of COVID-19 new increase since July. Therefore, many Regional Directorates of Health had no more empty hospital beds for reanimation nor hospitalisation, and military doctors and equipment are more solicited.

Consequently, some Moroccan Ministry of Health (MoH) consecutive decisions were taken. The decision n° 63 on August 05^th^, 2020 allowed to treat at home all asymptomatic COVID-19 confirmed only clinically, or after PCR confirmation for some suspected cases with confirmed contacts. The decision n°64 on August 13^th^, 2020 encouraged the implementation of a detection strategy by starting with a Point of Care Serological IgG/IgM test for all suspected patients. If positive (IgM positive) doing a RT-PCR and if negative nothing to do for the suspected patients. Then another decision n°69 on September 2^nd^, 2020 fostered the clinical role for defining symptomatic and asymptomatic cases and the deadlines to declare recovery after taking the standard treatment either at home or at the hospital. After that, a decision n°73 on September 16^th^, 2020 reorganised the first contact of the suspected COVID-19 at the nearest primary health centre where the PCR samples were allowed again to be done and referred to the public laboratory for getting a quick result. A decision n° 76 on September 26^th^, 2020 allowed all private laboratories to do RT PCR and Serological rapid test confirmation of SARS-CoV2 without any restrictions for who can be tested or not. Thus, everyone who can pay 60 to 70 $US as unitary molecular test cost, could get within a theoretical 48 hours a confirmation result. Finally, the decision n°80 on October 15^th^, 2020 promoted the start of media campaign around Influenza vaccination for old persons more than 64 years old and children under 5yo, by invitation to take the vaccine from the private sector at 12 $US per unit and defined an objective to get a 60% of inductive immunity in a specific targeted population that the MoH will vaccine free of charge (all health professionals and health students in the public sector, all pregnant women at the last six months and all hemodialyzed persons followed in the public sector).

The study aim is to share alternative COVID-19 case management based on the available scientific evidence to decrease deaths within a short time.

## Methods

This article methodology describes the use of qualitative content analysis to analyse official documents published about the COVID-19 case management in Morocco. The formal guideline and checklist of such research design are under development in EQUATOR Network database, as a new structure for quality improvement reports. The main steps cited are the brief description of context, key measures for improvement from one side (provider or government), what would constitute improvement from a different side (expert practices, patients or scientific evidence), analysis and interpretation, to reach a proposition of a strategy for change[3].

Then, the analysis method adopted is well known in social sciences and newly more used for health policy topics. We followed mainly the method described in *Hall&Steiner 2020* article[4]. This model was associated with the EQUATOR model, and multiple models suggesting the implementation of a modified strategy after decision analysis[5]. Therefore, the combined final model content analysis involves five steps:

- Presenting the legal or official documents
- Identifying the policy themes via a qualitative inductive reading of documents
- Describing the quantitative part of the policy parameters and trends
- Evaluating the spectrum of themes by qualitative deductive comparison to expert recommendations (based on the available published evidence)
- Suggesting a new approach to enhance the overall policy

The identification of the policy themes based on the six cited MoH decisions were organised by keywords and associated with the questions and hypotheses that policymakers consider. Then, **Figure1a**, in **supporting information**, is the generated framework of the first step.

For the first part, the parameters of COVID-19 in Morocco are notified by the number of new confirmed cases, new recovered persons, deaths and remaining active cases. The trends were analysed in Excel Microsoft Software and presented by figures. Secondly, a qualitative content analysis was done by NVivo (QSR International software), a software allowing to organize and document the reading by codes and sets. NVivo analysis gathered together the MoH decisions and the relevant articles issued from scientific evidence targeted by a parallel review[6].

Then the triangulation of the four last steps, themes selected from the MoH decisions, quantitative parameters, synthesis of the available scientific evidence helped to present the results as a coding tree **(Figure1b)**.

**Figure 1b:**
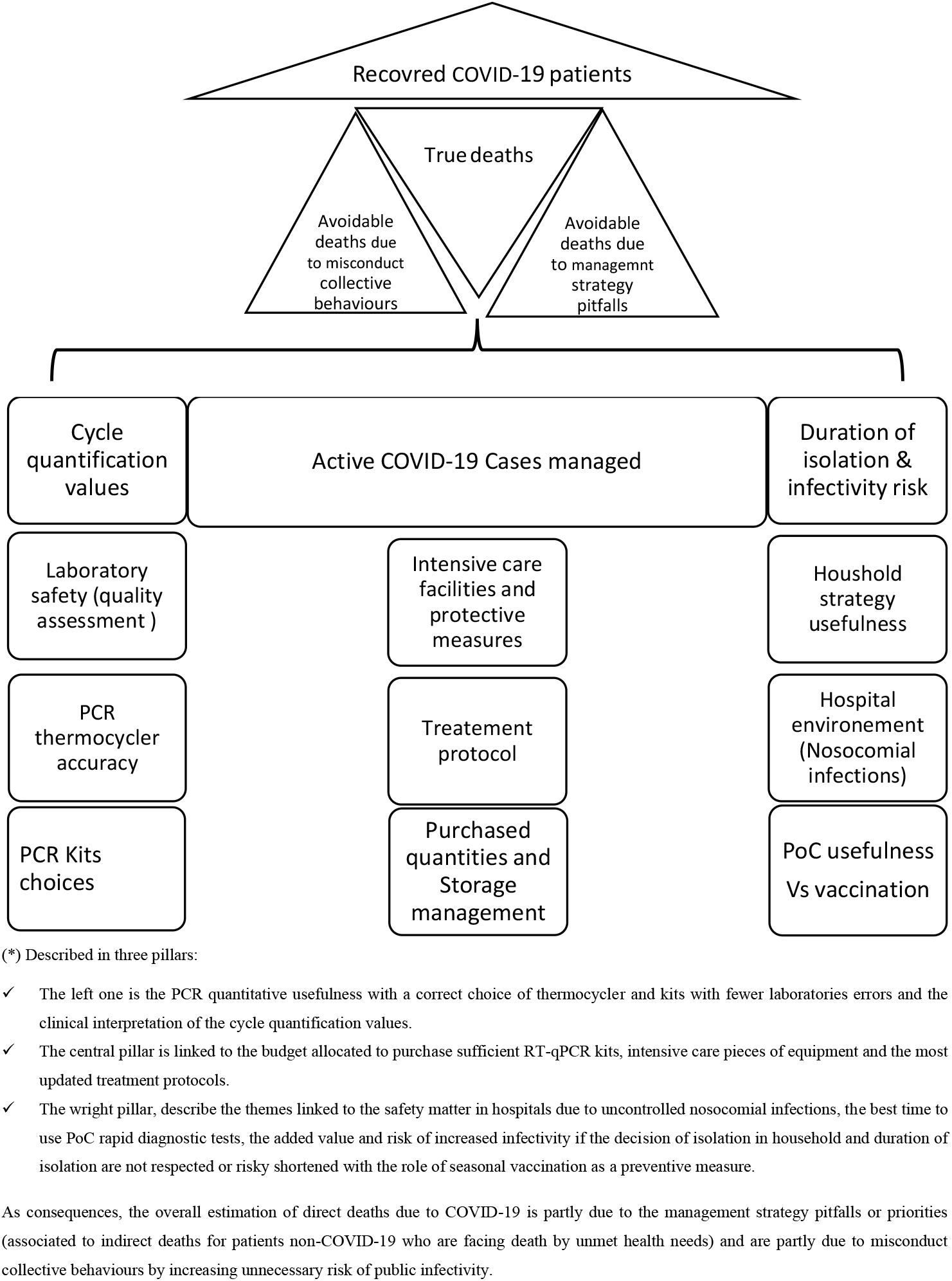
The NVivo thematic coding tree of COVID-19 binary outcomes and the influencing weighed strategic managerial decisions*.

The coding tree was divided into the COVID-19 primary outcomes (deaths or recovering states), and the keywords included in the MoH strategic decisions and hypothesis.

## Results

Twenty articles from the review were selected as presenting valuable evidence that confronts the policy themes. This section summarises the main themes gathered from the coding tree:

### I. COVID-19 parameters in Morocco and the lethality assessment

Morocco, with its 36 Million citizens, noticed the first imported case, on March 2^nd^, 2020. Then, the spread of COVID-19 cases in all the twelve administrative Moroccan regions raised many individual and collective cases. The metropolitans four cities (Casablanca the biggest one, Marrakech, Tangier, and Fez) had almost 70% of the cases. Unfortunately, a considerable increase is reported during August with around 20 to 40 deaths per day (e.g., On August 31 there was 62590 confirmed cumulative cases and 1141 cumulative deaths). However, during September, only Casablanca and Marrakech sustained a high level of confirmed cases, but the overall deaths remained stable between 40 ± 07 per day (e.g., On September 30 there was 102715 confirmed cumulative cases and 2194 cumulative deaths; unfortunately on October 31 the confirmed cumulative cases were 219084 with 3695 cumulative deaths).

The net lethality ratio is the total number of deaths due to COVID-19 divided by the overall confirmed COVID-19 cases. This ratio moves slowly around 1,7% and does not add any useful analysis **(Figure2)**. Thus, COVID-19 lethality could be presented with standardisation to a minimal number of the general population. (The days’ tests do no go further than 23000 tests per day for technical reasons, even by inviting all private laboratories to do SARS-CoV2 tests. The control strategy does not target mass enrolment of all suspected COVID-19 cases). As shown in **Figure3**, the standardised death ratio is expected to increase from less than ten deaths per one Million habitants to be more than 220 deaths per one Million citizens by the end of the year with an estimation around 7700 lives lost.

**Figure 2:**
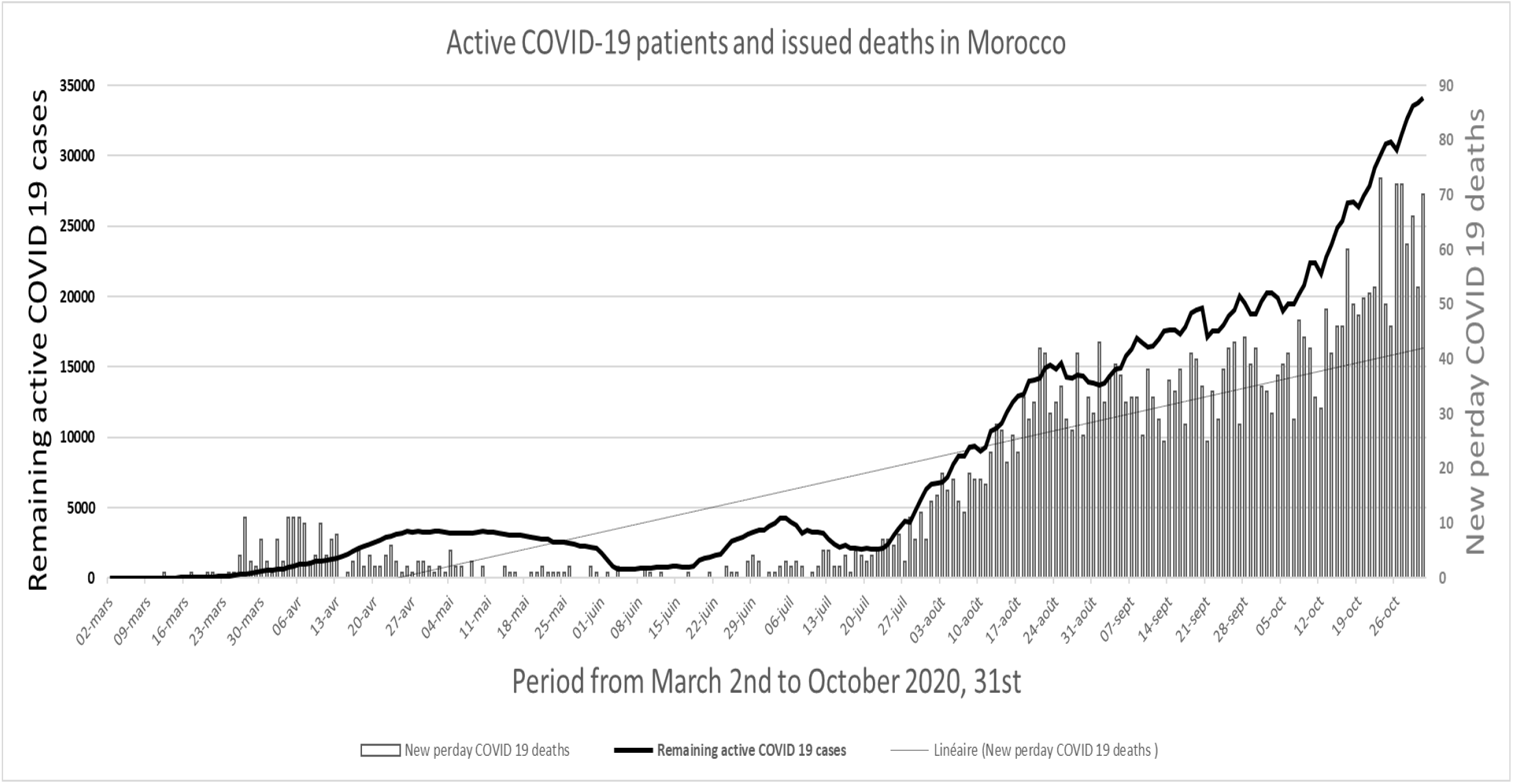
Active COVID-19 patients and issued deaths in Morocco between March and October 2020.

**Figure 3:**
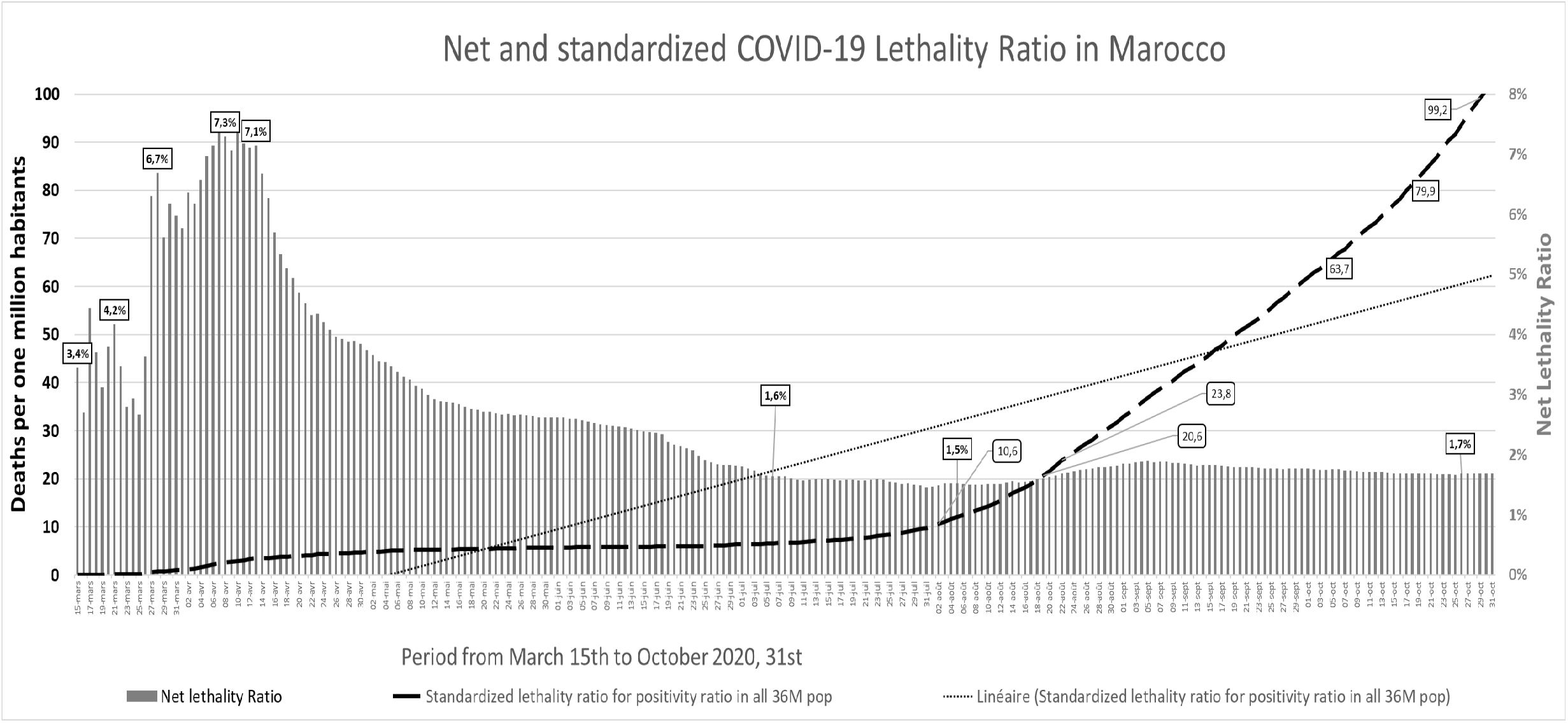
Net and standardised COVID-19 Lethality Ratio in Morocco from March to October 2020.

In other studies, the mortality rate of COVID-19 is commonly calculated comparing the numbers of patients who were discharged well recovered versus those who died by a timing endpoint (e.g. during the 30 past days) [7].

### II. Point of care (PoC) serology tests and vaccination as preventives measures

The IgG serodiagnosis rapid tests that could be used are not considered as the gold standard test due to their not satisfactory accuracy and are mostly used to show the seroconversion and the previous exposure to the virus after at least ten days from the onset of the symptoms. In Morocco, the rapid test trademark used showed in foreign studies a better sensitivity after 14 days from the onset of symptoms and raised some concerns about its usefulness for COVID-19 confirmation[8]. Controversially, the molecular test of the same trademark was reported as highly accurate in the USA[9]. More information about PoC assays accuracy for Detection SARS-CoV-2 become available, and none of the published studies considers the non-ELISA PoC ones as adapted for detection of suspected patients either symptomatic or asymptomatic, due to the high risk of false negatives especially during the beginning days from the onset[10–16]. While some countries, specified and updated a pre-defined list of the approved Serological COVID-19 tests, based on the evaluation of their accuracy[6]. Indeed, some SARS-CoV-2 IgG antibody ELISA assay has 100% sensitivity and 95% specificity after 14 days from the onset[17].

To face the winter season, and the emergence of seasonal influenza in the same population threatened by COVID-19, many international laboratories are developing a new generation of PoC allowing detection of both viruses in the same test, unfortunately, the accuracy assessment would not be available before the end of the winter in the Northern hemisphere.

Moreover, for this year, the anti-flu vaccine is expected to be a quadrivalent influenza vaccine containing one strain from each B lineage in addition to H1N1 and H3N2 strains[18]. The intention to be vaccinated by seasonal influenzas increase with the COVID-19 risk perception and increase of age in the UK[19]. Then, the yearly quantities should be increased. Another study from Brazil explains the presence of an association between the inactivated trivalent influenza vaccine and lower mortality among Covid-19 patients; such study results are available due to the seasonal vaccination in the southern hemisphere done in past April and May in concomitance with the COVID-19 emergence[20]. However, the results could be taken with more caution due to the possible presence of a herd immunity effect that allowed less mortality as reported in another previous study from a small town located in Brazil[21].

Furthermore, in vitro experiment, from multi European research collaboration, confirmed that quadrivalent inactivated influenza vaccine induces trained immunity responses against SARS-CoV-2 that enhance the COVID-19 protection[22]. Many previous studies demonstrated this vaccination benefice for pregnant women, and their newborns protected against influenza via passively acquired antibodies[23]. The quadrivalent vaccine immunogenicity and safety were confirmed for children aged 6-35 months and in older subjects aged 66-80 years[24]. The same well-tolerated seasonal influenza vaccine trademark is used in Morocco [25]. Additionally, based on the literature, all symptomatic or pre-symptomatic or asymptomatic COVID-19 patients should receive immediate seasonal influenza vaccination and the suitable period is between the second and twelve days from the onset to match with the best vaccination window’s opportunity for SARS-CoV-2 clearance[26].

### III. The RT-qPCR accuracy

For some scientists and health laboratory workers, the big challenge is that the PCR is not appropriate to detect virus infections, but it still used due to its ability to replicate DNA sequences by reverse transcriptase method. However, the amount of DNA obtained with the same RNA material can vary widely. As consequences, if the specificity of this technic is almost 95 %, its sensitivity is considered at least around 70%[27]

More, RNA extraction step represents a major bottleneck due to reagents shortage, cost, polling technic and time-consuming procedures[2,28]. Some clinical and testing errors (ineffective symptom screening, sampling errors, sample contamination,.) and analytical testing errors (insufficient sample, non-validated tests, instrument malfunction,.) compromise the results[11].

The Limit of Detection (LoD) depends on the trademark of the molecular assay. The low LoD may be attributable to technical deficiencies in the product’s manufacture[29]. In practice, the low sensitivity of the kit implies failing to identify many patients. Which means, at least up to 20% of all negative COVID-19 tests with RT-qPCR could be false negatives[30]. Similarly, a single negative test should not be used as a determinant clinical decision in patients [31,32].

In Morocco, the case management strategy was initially based on the PCR’ results to confirm the COVID-19’ positivity and the negative control results to declare recovering. However, the only information available at the operational level was the qualitative binary appreciation of this test (Positive, Or Negative). Indeed, some commercialised SARS-CoV-2 kits for RT-qPCR (the cheapest ones) are only qualitative, contrary to the fact that the “q” in “qPCR” stands for “quantitative". However, some manufacturers declare possible to use additional specific software’s to define the viral load by indirect calculations[31]. While, good laboratory practices emphasise that RT-qPCR should provide the viral load indirect and individual quantification, to allow to match with the required minimal information needed for results declaration[33].

### IV. The role of cycles quantification values

One of the public health benefices to investigate the cycles quantification values (Cq) (anciently named cycle threshold) of all affected persons, is to control the overall severity of the different periods of epidemy that happen in the country. As an example, Italy described the trend of the Cq values concerning different periods of the epidemic, showing a statistically significant increase in Cq values associated with a decrease in the percentage of affected samples[34]. For example, a person with a high Cq value tested early in the disease course might be or become infectious with a new lower Cq value. Then, the presence of more viral load is translated by a decrease in Cq value when the control test is done[34]. Under-treatment, the mean viral load decreased rapidly but could increase in one to three weeks [35].

However, COVID-19 recovered person is not easy to define. Each one who had no more symptoms needs two tests noticing negative results or close to the Cq >34 (do not have meaningful transmissibility of the disease)[36]. Indeed, Cq > 34 correspond to less than four viral copies in RT-qPCR device with 100% sensitivity[37,38]. A French team confirmed the strong correlation between successful isolation of SARS-CoV-2 in cell culture and Cq value of RT-qPCR targeting envelop gene and suggested that patients with Cq above 34 (Based on their PCR device used) are not contagious and can be exempted from hospital care or for strict confinement of the non-hospitalised patients[39]. While being sure that recovered patients are not in pre-relapse phase, or remain with infra clinical symptoms, and genes or spikes mutations are other scientific challenges[40,41].

### V. Household stay and asymptomatic cases management

The asymptomatic cases could be an essential source of contagion and need to be controlled biologically via virus nucleic acid testing[42]. Even if, the presence of RNA in RT-qPCR does not necessarily correlate with infectivity or transmission capacity, or at least is not yet easy to prove it scientifically[43]. However, studies published before July 2020 reported that the viral load detected was similar in both asymptomatic and the symptomatic patients, which prove the transmission potential of asymptomatic patients and minimally symptomatic ones [35,42,44],

Since August 01^st^, in Morocco, due to more severe cases needing intensive emergency cares and hospital beds, the asymptomatic patients or those with few and minim symptoms are invited to take the adjuvant medication and stay in their homes **(Figure1a)**. While, the evidence from China and USA show that households sites are the most favourable areas to spread the disease between 33% and 55% of the parents, partners, spouses and children were affected by their positive in-home person living with them. The odds ratio of the infectivity increases by seven to fifteen times if the residents in the household have some associated morbidities such diabetes or immunocompromised health conditions[45,46]. Ignorance, small habitations, less favourable social conditions, and inappropriate houses living conditions are the common risk factors for more inhouse infectivity.

### VI. The duration of isolation and infectivity risk

Substantial viral loads can be detected around day five of infection and decrease gradually based on the characteristics of the disease or the effective antiviral treatments taken[35,47]. In contrast, the virus load and transmission events start earlier two to three days before symptom onset[48]. While at the beginning of the pandemic, some studies 14 days of isolation after diagnosis stated to be sufficient to get negativity [49]. In contrast from Wuhan in China, the onset of the symptoms was linked to the percentage of positive results. This positivity declined from 100% in week one, to 66% in week three, and 32% in week four, to 5 and 0% in weeks five and six[36]. Then, the accuracy is correlated to the duration from the symptoms onsets, and the day 24^th^ is the mean date to get negativity[36]. Another study from Italy reported that the timeline of Cq value would be negative between 21 and 28 days[50]. Then, for both patients and health workers suggest a longer time of self or supervised isolation[51].

In contrast, the MoH assume two unchangeable hypotheses that become useless: the person is no longer transmitting virus ten days after symptom resolution and COVID-19 patient’s loss their infectivity after seven days under treatment. Consequently, the isolation should be increased from 14 to 28 days and balanced with the following parameters (the asymptomatic state, the overall duration from the beginning of the symptoms, the younger vs older, the time change of the cycles quantification).

### VII. Hospital environment and laboratory safety matters

Staphylococcus aureus and Candida Albicans that are frequent in the Moroccan population and are the primary nosocomial infections in the Moroccan hospitals lead to false-positive SARS-CoV-2 results by primers’ cross-reactivity. The co-infection with other viruses or bacteria create diagnostic confusion like for the cold virus (Influenza, Parainfluenza, Rhinovirus…)[52].

Moreover, nosocomial infections are sustained by the high level of contamination of air and surface by SARS-CoV-2 in hospital rooms. One study from Singapore hospitals reported more than 56% of environmental hospital rooms contamination and more than 66% of hospital surface contamination[53]. In contrast, the Cq value of contaminated rooms and non-contaminated rooms were equal to 25 and 33, respectively[53].

False positives results occur in a laboratory if reagents become contaminated, which is a significant concern about the testing volume during a pandemic. Indeed, the occurrence of contaminations of commercial primers of SARS-CoV-2 affects diagnostic specificity. Thus, the need to pre-test each batch of reagents before using in routine [54]. Then, COVID-19 diagnostic results should be reassessed systematically with the clinical or radiological patterns to validate the epidemiological basis and take individualised measures.

In Morocco, following the epidemiological increase of SARS-CoV-2, more self-demands about molecular tests arise. This test could be done theoretically within a price of zero, 55 to 150 US dollars, depending on the health coverage and the laboratory affiliation and the laboratory package promoted (Public, Army or private sectors). The actual process to do self-tests or tests based on the population demand remains not available in Morocco, even if this strategy is presented as less consuming of personal protective equipment[55]. Effectively, insufficient laboratories number able to perform molecular tests with safety conditions, a shortage and no standardisation of testing reagents and equipment create delays in testing with reduced effectiveness to control the direct first contacts and their secondary contacts in each new growth outbreak. Either, to face more demand pressure, laboratories must perform their own validation pool studies to determine the most efficient pool size[28,56].

## Discussion

Well-designed COVID-19 case management could be achievable by understanding some decisions should not be in opposition to scientific evidence nor by limiting the use of molecular test and nor by introducing PoC as the first diagnostic for recent suspected cases or contacts leading to false-negative non treated patients, and nor by starting treating in-home asymptomatic cases with a notification of recovering based only on clinical criteria **(e**.**g. from Morocco; Figure1a**, available as **supporting information)**. If not, those technical decisions could contribute to the spread of the COVID-19.

Moreover, the scientific development, and the emerging of influenza, create another environmental situation that needs a new generation of PoC that allow dual detection of SARS-CoV2 and Influenzas lineages. Thus, the use of the old generation of serological tests become reasonable after the 14^th^ day of the onset as diagnostic follow up of the treated patients.

Additionally, the importance of the cycles quantification (Cq) values as an essential parameter at the clinical and the diagnostic levels, is scientifically sustained by forthcoming review and another review that documented the Cq usefulness[6,57]. Therefore, the Moroccan MoH and similar such context countries are invited to purchase or define sensitive RT-qPCR thermocyclers standardised to all the laboratories sites; with periodic assessments of the commercial SARS-CoV-2 molecular kits to control and build a list of the most practical ones.

Furthermore, the scientific results’ interpretation will be facilitated by the future development of automatic case management software based on artificial intelligence laboratory information system that is meaningful for monitoring and evaluation[31,58]. With or without an intelligence result interpretation assistance, the **new model includes three phases of COVID-19 case management** that gather the specific information about the virus load dynamic of the patient’s categories and their evolutions:

**Phase 1- clinical interpretation of the expected viral load percentage:** a logical decision balanced between expectation of the viral load percentage progressing within weeks and the clinical state of each suspected patient as explained in **Table1**.

**Table 1.** Phase 1- The percentage to detect COVID-19 based on the natural evolution of the disease depending on patients clinical COVID 19 symptoms, clinical health assessment, age categorisation, comorbidities, immunocompetent health state and the onset day. **Clinical interpretation of the expected percentage of the viral load:** If the viral load percentage is estimated clinically (High) between 100% and 89%, One RT-qPCR will be done from sputum, pharynx, Saliva, or nasopharynx specimen. If negative, another immediate RT-qPCR will be done from nasopharynx specimen If the viral load percentage is estimated (Medium) at 66%, One RT-qPCR will be done from a nasopharynx specimen. If negative, another immediate RT-qPCR will be done from nasopharynx specimen If the viral load percentage is estimated clinically (low) between 32% and 05% just one RT-qPCR from the Nasopharynx should be done. If negative, surveillance for one week then IgM/IgG PoC rapid test

| Weeks after the COVID-19 first-day symptoms onset |  |  |  | One | Two | Three | Four | Five |
| --- | --- | --- | --- | --- | --- | --- | --- | --- |
| Percentage to detect the COVID-19 cases based on the Natural history of its viral load within weeks (*) |  |  |  | 100% | 89% | 66% | 32% | 05% |
| Clinical assessment + systematic seasonal vaccination (for each not vaccinated patient**) | For an asymptomatic young person (Less than 45 years old) without known chronic comorbidities | Discovered hazardously | Unknown (hypothesis of three days before the date of the first positive PCR) | 1/9 | 2/9 | 3/9 | 2/9 | 1/9 |
|  |  | Discovered by investigation of a positive contact of a previously confirmed case |  | 1/9 | 2/9 | 3/9 | 2/9 | 1/9 |
|  | For the symptomatic immunocompetent young person | with confusing suggestive clinical symptoms | Not well defined (three days before the best onset date remembered) | 2/8 | 3/8 | 2/8 | 1/8 |  |
|  |  | With intense suggestive clinical symptoms | Easy to remember | 3/6 | 2/6 | 1/6 |  |  |
|  | For a person at any age with chronic comorbidities | with confusing suggestive clinical symptoms | Not well defined (three days before the best onset date remembered) | 1/4 | 2/4 | 1/4 |  |  |
|  |  | With intense suggestive clinical symptoms | Easy to remember | 2/8 | 3/8 | 2/8 | 1/8 |  |
|  | For an older person with any health state or any patient with immunocompromised state | with confusing suggestive clinical symptoms | Not well defined (three days before the best onset date remembered) | 2/8 | 3/8 | 2/8 | 1/8 |  |
|  |  | With intense suggestive clinical symptoms | Easy to remember | 3/6 | 2/6 | 1/6 |  |  |
|  |  |  | 1<sup>st</sup> day of COVID-19 onset | The ratio of the probability of each patient's clinical health conditions reparation at the first confirmation |  |  |  |  |

(\*) Majority of patients (Around 70%) got positive results of RT-PCR test for SARS-CoV-2 within three weeks after the onset of symptoms. The negative results of RT-PCR test for SARS-CoV-2 began dominant from week four after the onset of symptoms. Reinfection could be the explanation of positive cases while they got the previous two consecutive negative results. (Xiao AT, Tong YX, Zhang S. Profile of RT-PCR for SARS-CoV-2: a preliminary study from 56 COVID-19 patients. Clin Infect Dis 2020. \*\*)Systematic seasonal vaccination for all suspected patients aged > 6 months

**Phase 2**- **diagnostic tree allowing confirmation and control:** based on the cycles quantification range (Cq) for all affected population as described in **Figure4**.

**Figure 4:**
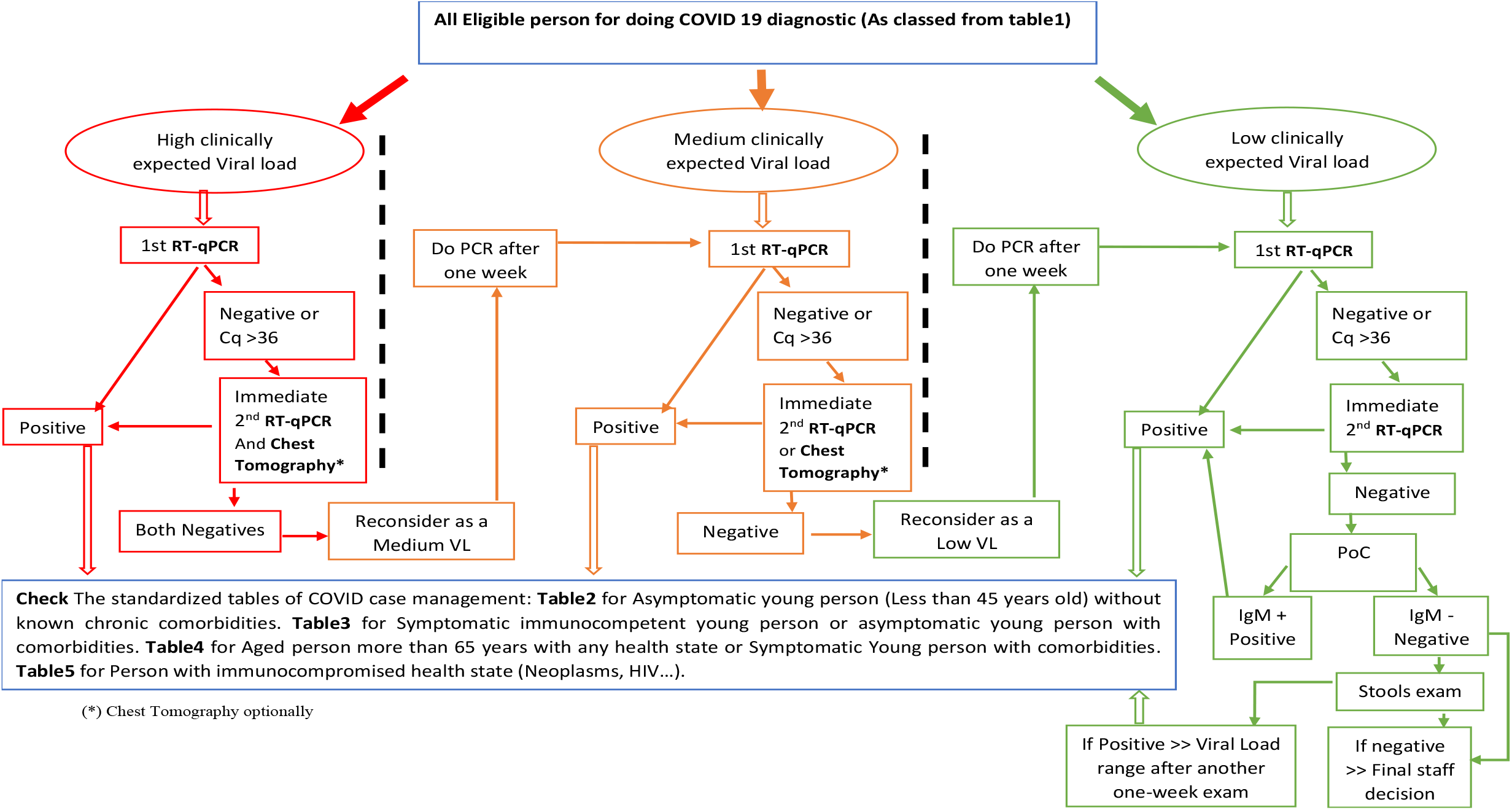
Phase 2- Molecular confirmation and diagnostic control decision tree.

**Phase 3- case management tables comparing patient category and Cq range**: Notify the Cq as three defined range, low, medium, and high. The standardised tables (**Tables 2**,**3**,**4**,**5** available as comprehensive **supporting information**) of COVID-19 case management target four different categories: Asymptomatic young person without known chronic comorbidities; Symptomatic immunocompetent young person or asymptomatic young person with comorbidities; Aged person with any health state or Symptomatic young person with comorbidities; Person with immunocompromised health state.

This study has some limits. Firstly, do not introduce the Radiology chest tomography (CT) results systematically as an individualised parameter of making the diagnostic, find the justification in the Moroccan context, the feasibility to get for each suspected person a laboratory test and a CT are heavy managerial tasks. However, all COVID-19 severe cases admitted in intensive unit care have CT. Moreover, a recent review showed that difference between the sensitivities of CT and RT-qPCR for SARS-CoV-2 is lower than previously thought. Secondly, not discussing any possible correlation between some routine blood laboratory exams and COVID-19 viral load is made, as there is no consensus about that[59,60]. Thirdly, the hypothesis explaining the protective link between the small percentage of the net lethality ratio in Morocco, and the mandatory BCG vaccination is not proved. Indeed, a retrospective cohort study found that BCG vaccination in childhood has no protective effects against COVID-19 in adulthood[61]. Another study from Sweden concludes to the absence of any protective effect against the COVID-19 in BCG vaccinated persons during infancy[62]. Fourthly, the effectiveness and time to get an adequate worldwide immunisation by a specific future COVID-19 vaccination, and it could be mandatory for all citizens, or only international travels remain questionable [63]. Finally, the MoH could mess a better cost-effective way to control this virus by taking for this winter season the mandatory vaccination against influenza that will be beneficial to limit indirectly COVID-19 deaths in the targeted population. That recommendation was not formulated, due to the end of the yearly expressed anti-flu vaccine needs and the official declaration to acquire the anti-COVID-19 vaccine as the main priority.

## Conclusions

Our work meets the same conclusion of other international researchers[36,57], RT-qPCR reported as a binary positive or negative result removes useful information that could inform clinical decision making or at least enhance it. This study proposes new case management to address the uncontrolled situation that could be adapted to local contexts and used by many other countries. On one side, massive seasonal vaccination to reach induced collective immunity level for all ordinary targeted population including the COVID-19 new suspected patients aged > 6 months, and on the other side, implement a mass molecular diagnostic control of COVID-19 as primary diagnostic intention. In comparison, local manufacturing of accurate PCR kits could be a cost-effective scenario to reach all diagnostic needs within the suspected population and its neighbourhood.

## Data Availability

The datasets used and analysed during the current study are available from the corresponding author on reasonable request.

## Abbreviations

COVID-19: Coronavirus disease 2019
SARS-CoV-2: Severe acute respiratory syndrome coronavirus 2
PCR: Polymerase chain reaction
RT-qPCR: Real-time quantitative PCR
RNA: Ribonucleic acid
DNA: Deoxy-ribonucleic acid
MoH: Ministry of Health
IgG/IgM: Immunoglobulin G/ Immunoglobulin M
PoC: Point of care
ELISA: Enzyme-linked immunosorbent assay
USA: United States of America
LoD: Limit of Detection
Cq: Cycles quantification value
CT: Chest tomography
BCG: Bacillus Calmette–Guérin

## Acknowledgements

Thanks to all providers, health professionals, civil and Military Institutions in Morocco for their commitment and hard work achievement to control as much as possible this disease. Solidarity expressed with all families, health professional and colleagues who faced the deadly consequences of COVID-19.

Ideas, results, and conclusions issued from this work and included in this manuscript, do not necessarily state the official position of the Ministry of Health or the author affiliation.

## Declarations

### Ethics approval and consent to participate

This paper does not contain any studies done by the authors involving human participants. It is based on a review of the literature and public statistical available data updated each day by the Moroccan Ministry of Health and available by the website link: http://www.covidmaroc.ma

### Source of Financing

No funding source supported the study

## Supporting information

### Tables and Figure available online-only

**Additional file Figure1a:**
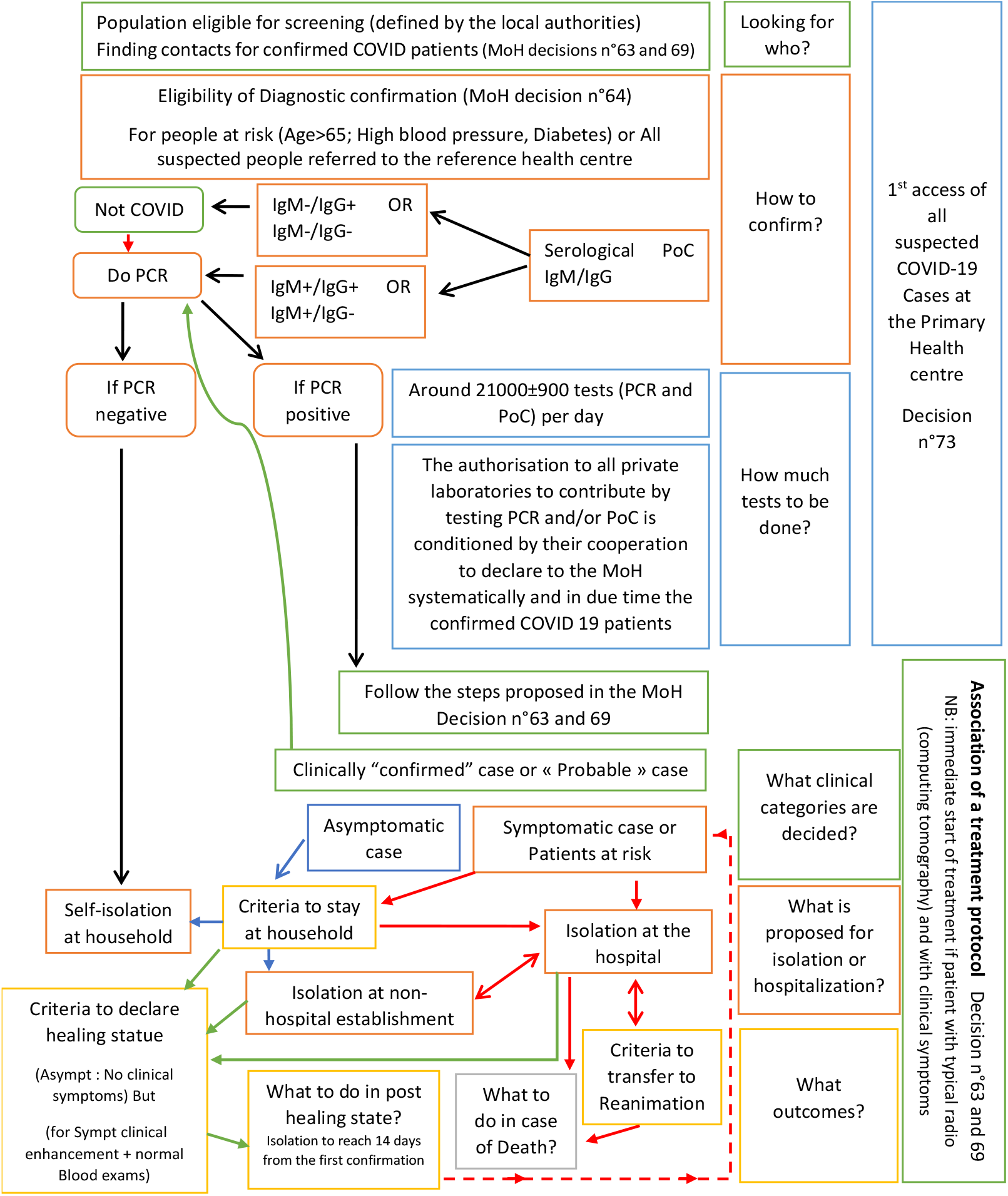
Framework of the proposed Moroccan COVID-19 case management based on the August and September MoH decisions (This framework do not include the implemented general rules of physical distancing, preventive gestures, hygiene, and mask utilisation permanently advised nor the vaccination plan which effectively start in November 2020).

**Additional file Table 2.** **Phase 3-** COVID19 case management of **Asymptomatic young person (Less than 45 years old) without known chronic comorbidities**, with regard of RT-qPCR (diagnostic result vs control result) based on the interpretation of the Cq range progress:

| Diagnostic process based on the Cq range of the PCR diagnostic result and the PCR control result (one week after) |  | Diagnostic interpretation based on the cycle quantification (or threshold cycle) |  |  |  |  |
| --- | --- | --- | --- | --- | --- | --- |
| | | 1<sup>st</sup> Cq between [16-33] then control test Cq stays between [16-33] (one week after) | 1<sup>st</sup> Cq between [16-33] then control test Cq [34-37](one week after) | 1<sup>st</sup> test Cq [34-37[ then control test Cq [34-37[ (one week after) | 1<sup>st</sup> test Cq [34-37[ then control test Cq between [16-33] (one week after) | 1<sup>st</sup> test negative (or extremely low positivity with Cq ≥ 37) and stay the same (one week after) |
| Cq qualitative variation |  | High or medium to high or medium | High or medium to low | Low to low | Low to high or medium | Stay Negative (Background) or extremely low |
| Asymptomatic young person (Less than 45 years old) without known chronic comorbidities | High clinical viral load presumption | Very high risk of transmission and complications during the 1<sup>st</sup> week. Isolation inside the hospital and monitoring for reanimation | High risk of transmission during the 1<sup>st</sup> week. Isolation inside the hospital then release | Probable New affection onset<br>Initially isolation into the hospital then-No isolation for no risk | Probable New affection onset<br>Initially isolation into the hospital then preparation to Intensive Emergency Care for high risk | After the one week, should consider the patient with a Medium clinical viral load presumption (follow the row below) |
|  | Medium clinical viral load presumption | High risk of transmission isolation in hospital | Medium risk of transmission during the 1<sup>st</sup>-week isolation outside the hospital then release | Probable New affection onset<br>Initially isolation outside hospital then-No isolation for no risk | Probable New affection onset<br>Initially isolation outside the hospital then Isolation in Hospital | After the one week, should consider the patient with a Low clinical viral load presumption (follow the row below) |
|  | Low clinical viral load presumption | Medium risk of transmission<br>Isolation outside then isolation in hospital | Low risk of transmission during the 1<sup>st</sup>-week isolation outside the hospital then release | No risk of transmission No isolation Declaration of the negativity of the case and declaration as a healed case (one month after with a negative PoC result) | New affection onset<br>No isolation initially then isolation in hospital | No risk of transmission and no isolation Declaration of the negativity of the case and declaration as a healed case (one month after with a negative PoC result) |

**NB:** The notification should be systematic for all positive results by reporting the Cq values. (Consecutive two negative tests at the control time repeated twice after 14 and 28 days or after 20 and 28 days could be justified in the current context to declare recovery if both negatives)

**Additional file Table 3.** **Phase 3-** COVID-19 case management of **Symptomatic immunocompetent young person or asymptomatic young person with comorbidities**, with regard of RT-qPCR (diagnostic result vs control result) based on the interpretation of the Cq range progress:

| Diagnostic process based on the Cq range of the PCR diagnostic result and the PCR control result (one week after) |  | Diagnostic interpretation based on the cycle quantification (or threshold cycle) |  |  |  |  |
| --- | --- | --- | --- | --- | --- | --- |
| | | 1<sup>st</sup> Cq between [16-33] then control test Cq stays between [16-33] (one week after) | 1<sup>st</sup> Cq between [16-33] then control test Cq [34-37](one week after) | 1<sup>st</sup> test Cq [34-37[ then control test Cq [34-37[ (one week after) | 1<sup>st</sup> test Cq [34-37[ then control test Cq between [16-33] (one week after) | 1<sup>st</sup> test negative (or extremely low positivity with Cq ≥ 37) and stay the same (one week after) |
| Cq qualitative variation |  | High or medium to high or medium | High or medium to low | Low to low | Low to high or medium | Stay Negative (Background) or extremely low |
| The symptomatic immunocompetent young person or Asymptomatic young person with comorbidities | High clinical viral load presumption | Very high risk of transmission and complications during the 1<sup>st</sup> week. Isolation inside the hospital and monitoring for reanimation | High risk of transmission during the 1<sup>st</sup> week. Isolation inside the hospital then release | Probable New affection onset<br>Initially isolation into the hospital Then, No isolation for no risk | Probable New affection onset<br>Initially isolation into the hospital then preparation to Intensive Emergency Care for high risk | After the one week, should consider the patient with a Medium clinical viral load presumption (follow the row below) |
|  | Medium clinical viral load presumption | High risk of transmission<br>Isolation in Hospital | Medium risk of transmission during the 1<sup>st</sup>-week isolation outside the hospital then release | Probable New affection onset<br>Initially isolation into the hospital Then, No isolation for no risk | Probable New affection onset<br>Initially isolation outside the hospital then Isolation in Hospital | After the one week, should consider the patient with a Low clinical viral load presumption (follow the row below) |
|  | Low clinical viral load presumption | Medium risk of transmission<br>Isolation outside then Isolation in Hospital | Low risk of transmission during the 1<sup>st</sup>-week isolation outside the hospital then release | No risk of transmission No isolation Then, declaration of the negativity of the case two months after with a negative PoC result | New affection onset<br>No isolation initially then isolation in hospital | No risk of transmission, No isolation. Then, declaration of the negativity of the case two months after with a negative PoC result |

**NB:** The notification should be systematic for all positive results by reporting the Cq values. (Consecutive two negative tests at the control time repeated twice after 14 and 28 days or after 20 and 28 days could be justified in the current context to declare recovery if both negatives)

**Additional file Table 4.** **Phase 3-** COVID-19 case management of **Aged person more than 65 years with any health state or Symptomatic Young person with comorbidities**, with regard of RT-qPCR (diagnostic result vs control result) based on the interpretation of the Cq range progress:

| Diagnostic process based on the Cq range of the PCR diagnostic result and the PCR control result (one week after) |  | Diagnostic interpretation based on the cycle quantification (or threshold cycle) |  |  |  |  |
| --- | --- | --- | --- | --- | --- | --- |
| | | 1<sup>st</sup> Cq between [16-33] then control test Cq stays between [16-33] (one week after) | 1<sup>st</sup> Cq between [16-33] then control test Cq [34-37] (one week after) | 1<sup>st</sup> test Cq [34-37] then control test Cq [34-37] (one week after) | 1<sup>st</sup> test Cq [34-37] then control test Cq between [16-33] (one week after) | 1<sup>st</sup> test negative (or extremely low positivity with Cq ≥ 37) and stay the same (one week after) |
| Cq qualitative variation |  | High or medium to high or medium | High or medium to low | Low to low | Low to high or medium | Stay Negative (Background) or extremely low |
| An aged person more than 65 years with any health state or Symptomatic Young person with any health state | High clinical viral load presumption | Very high risk of transmission and complications during the 1<sup>st</sup> week. Isolation inside the hospital and monitoring for reanimation | High risk of transmission during the 1<sup>st</sup> week. Isolation inside the hospital then release | Probable New affection onset<br>Initially Isolation inside hospital then-No isolation then confirmation of negativity by PoC after two months | Probable New affection onset<br>Initially isolation into the hospital then preparation to Intensive Emergency Care for high risk | After the one week, should consider the patient with a Medium clinical viral load presumption (follow the row below) |
|  | Medium clinical viral load presumption | High risk of transmission<br>Isolation in Hospital | Medium risk of transmission during the 1<sup>st</sup>-week isolation in hospital then release | Probable New affection onset<br>Initially Isolation outside hospital then-No isolation then confirmation of negativity by PoC after two months | Probable New affection onset<br>Isolation inside the hospital and monitoring for reanimation | After the one week, should consider the patient with a Low clinical viral load presumption (follow the row below) |
|  | Low clinical viral load presumption | Medium risk of transmission<br>Isolation in Hospital | Medium risk of transmission during the 1<sup>st</sup>-week isolation in hospital then release | No risk of transmission No isolation then confirmation of negativity by PoC after two months | New affection onset<br>Isolation inside the hospital and monitoring for reanimation | Declaration of the negativity of the case and declaration as a healed case (three months after with a negative PoC result) |

**NB:** The notification should be systematic for all positive results by reporting the Cq values. (Consecutive two negative tests at the control time repeated twice after 14 and 28 days or after 20 and 28 days could be justified in the current context to declare recovery if both negatives)

**Additional file Table 5.** **Phase 3-** COVID-19 case management of **a person with an immunocompromised health state**, with regard of RT-qPCR (diagnostic result vs control result) based on the interpretation of the Cq range progress:

| Diagnostic process based on the Cq range of the PCR diagnostic result and the PCR control result (one week after) |  | Diagnostic interpretation based on the cycle quantification (or threshold cycle) |  |  |  |  |
| --- | --- | --- | --- | --- | --- | --- |
| | | 1<sup>st</sup> test negative (or extremely low positivity with Cq ≥ 37) and stay the same (one week after) | 1<sup>st</sup> test Cq≥34 then control test Cq ≥ 34 (one week after) | 1<sup>st</sup> test negative (or extremely low positivity with Cq ≥ 37) and stay the same (one week after) | 1<sup>st</sup> Cq between 16 and 33 then control test Cq ≥ 34 | 1<sup>st</sup> test negative (or extremely low positivity with Cq ≥ 37) and stay the same (one week after) |
| Cq qualitative variation |  | High or medium to high or medium | High or medium to low | Low to low | Low to high or medium | Stay Negative (Background) or extremely low |
| A person with an immunocompromised health state (Neoplasms, HIV...) | High clinical viral load presumption | Very high risk of transmission and complications during the 1<sup>st</sup> week. Isolation inside the hospital and monitoring for reanimation | High risk of transmission during the 1<sup>st</sup> week. Isolation inside the hospital | Medium risk of transmission during the 1<sup>st</sup> week. Isolation inside the hospital | Very high risk of transmission and complications during the 1<sup>st</sup> week. Isolation inside the hospital and monitoring for reanimation | After the one week, should consider the patient with a Low clinical viral load presumption |
|  | Medium clinical viral load presumption | Very high risk of transmission and complications during the 1<sup>st</sup> week. Isolation inside the hospital and monitoring for reanimation | High risk of transmission during the 1<sup>st</sup> week. Isolation inside the hospital | Medium risk of transmission during the 1<sup>st</sup> week. Isolation inside the hospital | Very high risk of transmission and complications during the 1<sup>st</sup> week. Isolation inside the hospital and monitoring for reanimation | After the one week, should consider the patient with a Medium clinical viral load presumption |
|  | Low clinical viral load presumption | Very high risk of transmission and complications during the 1<sup>st</sup> week. Isolation inside the hospital | High risk of transmission during the 1<sup>st</sup> week. Isolation inside the hospital | Medium risk of transmission during the 1<sup>st</sup> week. Isolation inside the hospital | High risk of transmission during the 1<sup>st</sup> week. Isolation inside the hospital | Declaration of the negativity of the case and declaration as a healed case (six months after with negative Molecular results) |

**NB:** The notification should be systematic for all positive results by reporting the Cq values. (Consecutive two negative tests at the control time repeated twice after 14 and 28 days or after 20 and 28 days could be justified in the current context to declare recovery if both negatives)

**Additional file Table 6:** The 28 articles selected from the Minireview and used for triangulation in this policy model content analysis

| ID | Title | Journal | Year of Publication |
| --- | --- | --- | --- |
| 100 | A Novel Multiplex qRT-PCR Assay to Detect SARS-CoV-2 Infection: High Sensitivity and Increased Testing Capacity | Microorganisms | 2020 |
| 34 | Comparison of commercial realtime reverse transcription PCR assays for the detection of SARS-CoV-2 | Journal of Clinical Virology | 2020 |
| 84 | Comparison of SARS-CoV-2 detection from nasopharyngeal swab samples by the Roche Cobas 6800 SARS-CoV-2 test and a laboratory-developed real-time RT-PCR test | Journal of medical virology | 2020 |
| 149 | Detection of air and surface contamination by SARS-CoV-2 in hospital rooms of infected patients | Nature communications | 2020 |
| 85 | Evaluating the efficiency of specimen pooling for PCR-based detection of COVID-19 | Journal of medical virology | 2020 |
| 70 | Evaluation of a quantitative RT-PCR assay for the detection of the emerging coronavirus SARS-CoV-2 using a high throughput system | Euro surveillance | 2020 |
| 24 | Interpret with caution: An evaluation of the commercial AusDiagnostics versus in-house developed assays for the detection of SARS-CoV-2 virus | Journal of clinical virology | 2020 |
| 75 | Limits of Detection of 6 Approved RT-PCR Kits for the Novel SARS-Coronavirus-2 (SARS-CoV-2) | Clinical chemistry | 2020 |
| 64 | Optimisation of the CDC Protocol of Molecular Diagnosis of COVID-19 for Timely Diagnosis | Diagnostics | 2020 |
| 73 | Potential False-Negative Nucleic Acid Testing Results for Severe Acute Respiratory Syndrome Coronavirus 2 from Thermal Inactivation of Samples with Low Viral Loads | Clinical chemistry | 2020 |
| 6 | Dynamic change process of target genes by RT-PCR testing of SARS-Cov-2 during the course of a Coronavirus Disease 2019 patient | Clinica Chimica Acta journal | 2020 |
| 132 | The MIQE Guidelines: Minimum Information for Publication of Quantitative Real-Time PCR Experiments | Clinical Chemistry | 2020 |
| 99 | Viral RNA load as determined by cell culture as a management tool for discharge of SARS-CoV-2 patients from infectious disease wards | European journal of clinical microbiology & infectious diseases | 2020 |
| 4 | **Virological surveillance of SARS-CoV-2 in an Italian northern area: comparison of Real-Time RT PCR cycle threshold (Ct) values in three epidemic periods** | **Acta bio-medica** | **2020** |
| 20 | **Evidence from two cases of asymptomatic infection with SARS-CoV-2: Are 14 days of isolation sufficient?** | **International Journal of infectious diseases** | **2020** |
| 104 | **Laboratory information system requirements to manage the COVID-19 pandemic: A report from the Belgian national reference testing centre** | **Journal of the American Medical Informatics Association: JAMIA** | **2020** |
| 140 | **SARS-CoV-2 Viral Load in Upper Respiratory Specimens of Infected Patients** | **New England Journal of Medicine** | **2020** |
| 138 | **Temporal dynamics in viral shedding and transmissibility of COVID-19** | **Nature Medicine** | **2020** |
| 143 | **Test Agreement between Roche Cobas 6800 and Cepheid GeneXpert Xpress SARS-CoV-2 Assays at High Cycle Threshold Ranges** | **Journal of clinical microbiology** | **2020** |
| 74 | **Time Length of Negativization and Cycle Threshold Values in 182 Healthcare Workers with Covid-19 in Milan, Italy: An Observational Cohort Study** | **International journal of environmental research and public health** | **2020** |
| 109 | **Viral dynamics in asymptomatic patients with COVID-19** | **International Journal of infectious diseases** | **2020** |
| 46 | **Assessment of Specimen Pooling to Conserve SARS-CoV-2 Testing Resources** | **American journal of clinical pathology** | **2020** |
| 139 | **Chronological Changes of Viral Shedding in Adult Inpatients With COVID-19 in Wuhan, China** | **Clinical Infectious Diseases** | **2020** |
| 14 | **Occult SARS-CoV-2 infection; a possible hypothesis for viral relapse** | **Medical hypotheses** | **2020** |
| 28 | **Self-collection: An appropriate alternative during the SARS-CoV-2 pandemic** | **Journal of clinical virology** | **2020** |
| 10 | **Tracking Changes in SARS-CoV-2 Spike: Evidence that D614G Increases Infectivity of the COVID-19 Virus** | **Cell** | **2020** |
| 32 | **Validation and verification of the Abbott RealTime SARS-CoV-2 assay analytical and clinical performance** | **Journal of clinical virology** | **2020** |
| 21 | **Viral kinetics of SARS-CoV-2 in asymptomatic carriers and presymptomatic patients** | **International Journal of infectious diseases** | **2020** |

**The scanned copies of the original MoH decisions** cited are available on request to the corresponding author and Moroccan Health authorities.

✓ PDF file of the Moroccan MoH decision n° 63 on August 05^th^, 2020
✓ PDF file of the Moroccan MoH decision n° 64 on August 13^th^, 2020
✓ PDF file of the Moroccan MoH decision n° 69 on September 2^nd^, 2020
✓ PDF file of the Moroccan MoH decision n° 73 on September 16^th^, 2020
✓ PDF file of the Moroccan MoH decision n° 76 on September 26^th^, 2020
✓ PDF file of the Moroccan MoH decision n° 80 on October 15^th^, 2020

## References

1 Chen C-J, Hsieh L-L, Lin S-K, et al. Optimization of the CDC Protocol of Molecular Diagnosis of COVID-19 for Timely Diagnosis. Diagnostics (Basel, Switzerland) 2020;10. doi:10.3390/diagnostics10050333

2 Song F, Zhang X, Zha Y, et al. COVID-19: Recommended sampling sites at different stages of the disease. J Med Virol Published Online First: 2020. doi:10.1002/jmv.25892

3 Moss F, Thompson R. A new structure for quality improvement reports. The EQUATOR-Network. https://www.equator-network.org/reporting-guidelines/a-new-structure-for-quality-improvement-reports/

4 Hall DM, Steiner R. Policy content analysis: Qualitative method for analyzing sub-national insect pollinator legislation. MethodsX 2020;7:100787. doi:10.1016/j.mex.2020.100787

5 Shea BK, Runge MC, Pannell D, et al. Harnessing multiple models for outbreak management. Science (80-.). 2020;368:577–9. doi:10.1126/science.abb9934

6 Bennis I, Nadia B. Cycles quantification value to enhance the SARS-CoV-2 Molecular diagnostics: a Minireview. Forthcoming peer-reviewed manuscript in 2021.

7 Han Z, Battaglia F, Terlecky SR. Discharged COVID-19 Patients Testing Positive Again for SARS-CoV-2 RNA: A Minireview of Published Studies from China. J Med Virol Published Online First: July 2020. doi:10.1002/jmv.26250

8 Chew KL, Tan SS, Saw S, et al. Clinical evaluation of serological IgG antibody response on the Abbott Architect for established SARS-CoV-2 infection. Clin Microbiol Infect Published Online First: 2020. doi:10.1016/j.cmi.2020.05.036

9 Degli-Angeli E, Dragavon J, Huang M-L, et al. Validation and verification of the Abbott RealTime SARS-CoV-2 assay analytical and clinical performance. J Clin Virol 2020;129:104474. doi:10.1016/j.jcv.2020.104474

10 Rahman H, Carter I, Basile K, et al. Interpret with caution: An evaluation of the commercial AusDiagnostics versus in-house developed assays for the detection of SARS-CoV-2 virus. J Clin Virol 2020;127:104374. doi:10.1016/j.jcv.2020.104374

11 Shyu D, Dorroh J, Holtmeyer C, et al. Laboratory Tests for COVID-19: A Review of Peer-Reviewed Publications and Implications for Clinical Use. Mo Med 2020;117:184–95.

12 Mathuria JP, Yadav R, Rajkumar. Laboratory diagnosis of SARS-CoV-2 - A review of current methods. J Infect Public Health 2020;13:901–5. doi:10.1016/j.jiph.2020.06.005

13 Oliveira BA, Oliveira LC de, Sabino EC, et al. SARS-CoV-2 and the COVID-19 disease: a mini review on diagnostic methods. Rev Inst Med Trop Sao Paulo 2020;62:e44. doi:10.1590/S1678-9946202062044

14 Abduljalil JM. Laboratory diagnosis of SARS-CoV-2: available approaches and limitations. New microbes new Infect 2020;36:100713. doi:10.1016/j.nmni.2020.100713

15 Esbin MN, Whitney ON, Chong S, et al. Overcoming the bottleneck to widespread testing: a rapid review of nucleic acid testing approaches for COVID-19 detection. RNA 2020;26:771–83. doi:10.1261/rna.076232.120

16 Harahwa T, Lai Yau TH, Lim-Cooke M-S, et al. The optimal diagnostic methods for COVID-19. Diagnosis 2020;0. doi:10.1515/dx-2020-0058

17 Kohmer N, Westhaus S, Rühl C, et al. Clinical performance of different SARS-CoV-2 IgG antibody tests. J Med Virol 2020;:pjmv.26145. doi:10.1002/jmv.26145

18 Rudenko L, Kiseleva I, Krutikova E, et al. Rationale for vaccination with trivalent or quadrivalent live attenuated influenza vaccines: Protective vaccine efficacy in the ferret model. PLoS One 2018;13. doi:10.1371/journal.pone.0208028

19 Bachtiger P, Adamson A, Chow J-J, et al. The Impact of the Covid-19 Pandemic on Uptake of Influenza Vaccine: A UK-Wide Observational Study. medRxiv 2020;:2020.10.01.20205385. doi:10.1101/2020.10.01.20205385

20 Fink G, Orlova-Fink N, Schindler T, et al. Inactivated trivalent influenza vaccine is associated with lower mortality among Covid-19 patients in Brazil. medRxiv 2020;:2020.06.29.20142505. doi:10.1101/2020.06.29.20142505

21 Buss LF, Prete Jr CA, Abrahim CM, et al. COVID-19 herd immunity in the Brazilian Amazon. medRxiv 2020;:2020.09.16.20194787. doi:10.1101/2020.09.16.20194787

22 Debisarun PA, Struycken P, Domínguez-Andrés J, et al. The effect of influenza vaccination on trained immunity: impact on COVID-19. medRxiv 2020;:2020.10.14.20212498. doi:10.1101/2020.10.14.20212498

23 Vesikari T, Virta M, Heinonen S, et al. Immunogenicity and safety of a quadrivalent inactivated influenza vaccine in pregnant women: a randomized, observer-blind trial. Hum Vaccines Immunother 2020;16:623–9. doi:10.1080/21645515.2019.1667202

24 Montomoli E, Torelli A, Manini I, et al. Immunogenicity and safety of the new inactivated quadrivalent influenza vaccine vaxigrip tetra: Preliminary results in children ≥6 months and older adults. Vaccines. 2018;6. doi:10.3390/vaccines6010014

25 Gresset-Bourgeois V, Leventhal PS, Pepin S, et al. Quadrivalent inactivated influenza vaccine (VaxigripTetra^™^). Expert Rev Vaccines 2018;17:1–11. doi:10.1080/14760584.2018.1407650

26 Jeyanathan M, Afkhami S, Smaill F, et al. Immunological considerations for COVID-19 vaccine strategies. Nat. Rev. Immunol. 2020;20:615–32. doi:10.1038/s41577-020-00434-6

27 Arevalo-Rodriguez I, Buitrago-Garcia D, Simancas-Racines D, et al. False-negative results of initial RT-PCR assays for COVID-19: A systematic review. Published Online First: 2020. doi:10.1101/2020.04.16.20066787

28 Wacharapluesadee S, Kaewpom T, Ampoot W, et al. Evaluating the efficiency of specimen pooling for PCR-based detection of COVID-19. J Med Virol Published Online First: May 2020. doi:10.1002/jmv.26005

29 Wang X, Yao H, Xu X, et al. Limits of Detection of 6 Approved RT-PCR Kits for the Novel SARS-Coronavirus-2 (SARS-CoV-2). Clin. Chem. 2020;66:977–9. doi:10.1093/clinchem/hvaa099

30 Pan Y, Long L, Zhang D, et al. Potential False-Negative Nucleic Acid Testing Results for Severe Acute Respiratory Syndrome Coronavirus 2 from Thermal Inactivation of Samples with Low Viral Loads. Clin Chem 2020;66:794–801. doi:10.1093/clinchem/hvaa091

31 Pujadas E, Ibeh N, Hernandez MM, et al. Comparison of SARS-CoV-2 detection from nasopharyngeal swab samples by the Roche cobas 6800 SARS-CoV-2 test and a laboratory-developed real-time RT-PCR test. J Med Virol Published Online First: May 2020. doi:10.1002/jmv.25988

32 Lv D-F, Ying Q-M, Weng Y-S, et al. Dynamic change process of target genes by RT-PCR testing of SARS-Cov-2 during the course of a Coronavirus Disease 2019 patient. Clin. Chim. Acta. 2020;506:172–5. doi:10.1016/j.cca.2020.03.032

33 Bustin SA, Benes V, Garson JA, et al. The MIQE guidelines: Minimum information for publication of quantitative real-time PCR experiments. Clin Chem 2009;55:611–22. doi:10.1373/clinchem.2008.112797

34 Veronesi L, Colucci ME, Pasquarella C, et al. Virological surveillance of SARS-CoV-2 in an Italian northern area: comparison of Real Time RT PCR cycle threshold (Ct) values in three epidemic periods. Acta Biomed 2020;91:19–21. doi:10.23750/abm.v91i9-S.10138

35 Huang J-T, Ran R-X, Lv Z-H, et al. Chronological Changes of Viral Shedding in Adult Inpatients with COVID-19 in Wuhan, China. Clin Infect Dis Published Online First: 23 May 2020. doi:10.1093/cid/ciaa631

36 Tom MR, Mina MJ. To Interpret the SARS-CoV-2 Test, Consider the Cycle Threshold Value. Clin Infect Dis Published Online First: 2020. doi:10.1093/cid/ciaa619

37 Petrillo S, Carrà G, Bottino P, et al. A Novel Multiplex qRT-PCR Assay to Detect SARS-CoV-2 Infection: High Sensitivity and Increased Testing Capacity. Microorganisms 2020;8. doi:10.3390/microorganisms8071064

38 Yoon JG, Yoon J, Song JY, et al. Clinical Significance of a High SARS-CoV-2 Viral Load in the Saliva. J Korean Med Sci 2020;35:e195. doi:10.3346/jkms.2020.35.e195

39 La Scola B, Le Bideau M, Andreani J, et al. Viral RNA load as determined by cell culture as a management tool for discharge of SARS-CoV-2 patients from infectious disease wards. Eur J Clin Microbiol Infect Dis 2020;39:1059–61. doi:10.1007/s10096-020-03913-9

40 Korber B, Fischer WM, Gnanakaran S, et al. Tracking Changes in SARS-CoV-2 Spike: Evidence that D614G Increases Infectivity of the COVID-19 Virus. Cell Published Online First: July 2020. doi:10.1016/j.cell.2020.06.043

41 Elberry MH, Ahmed H. Occult SARS-CoV-2 infection; a possible hypothesis for viral relapse. Med. Hypotheses. 2020;144:109980. doi:10.1016/j.mehy.2020.109980

42 Zhou R, Li F, Chen F, et al. Viral dynamics in asymptomatic patients with COVID-19. Int J Infect Dis 2020;96:288–90. doi:10.1016/j.ijid.2020.05.030

43 Iglói Z, Leven M, Abdel-Karem Abou-Nouar Z, et al. Comparison of commercial realtime reverse transcription PCR assays for the detection of SARS-CoV-2. J Clin Virol 2020;129:104510. doi:10.1016/j.jcv.2020.104510

44 Zou L, Ruan F, Huang M, et al. SARS-CoV-2 viral load in upper respiratory specimens of infected patients. N. Engl. J. Med. 2020;382:1177–9. doi:10.1056/NEJMc2001737

45 Lewis NM, Chu VT, Ye D, et al. Household Transmission of SARS-CoV-2 in the United States. Clin Infect Dis Published Online First: 2020. doi:10.1093/cid/ciaa1166/5893024

46 Wang Z, Ma W, Zheng X, et al. Household transmission of SARS-CoV-2 ☆. J Infect 2020;81:179–82. doi:10.1016/j.jinf.2020.03.040

47 Broder K, Babiker A, Myers C, et al. Test Agreement between Roche Cobas 6800 and Cepheid GeneXpert Xpress SARS-CoV-2 Assays at High Cycle Threshold Ranges. J. Clin. Microbiol. 2020;58. doi:10.1128/JCM.01187-20

48 He X, Lau EHY, Wu P, et al. Temporal dynamics in viral shedding and transmissibility of COVID-19. Nat Med 2020;26:672–5. doi:10.1038/s41591-020-0869-5

49 Kim SE, Jeong HS, Yu Y, et al. Viral kinetics of SARS-CoV-2 in asymptomatic carriers and presymptomatic patients. Int J Infect Dis 2020;95:441–3. doi:10.1016/j.ijid.2020.04.083

50 Cariani L, Orena BS, Ambrogi F, et al. Time Length of Negativization and Cycle Threshold Values in 182 Healthcare Workers with Covid-19 in Milan, Italy: An Observational Cohort Study. Int J Environ Res Public Health 2020;17. doi:10.3390/ijerph17155313

51 Wan R, Mao Z-Q, He L-Y, et al. Evidence from two cases of asymptomatic infection with SARS-CoV-2: Are 14 days of isolation sufficient? Int J Infect Dis 2020;95:174–5. doi:10.1016/j.ijid.2020.03.041

52 Pfefferle S, Reucher S, Nörz D, et al. Evaluation of a quantitative RT-PCR assay for the detection of the emerging coronavirus SARS-CoV-2 using a high throughput system. Euro Surveill 2020;25. doi:10.2807/1560-7917.ES.2020.25.9.2000152

53 Chia PY, Coleman KK, Tan YK, et al. Detection of air and surface contamination by SARS-CoV-2 in hospital rooms of infected patients. Nat Commun 2020;11:2800. doi:10.1038/s41467-020-16670-2

54 Wernike K, Keller M, Conraths FJ, et al. Pitfalls in SARS-CoV-2 PCR diagnostics. Transbound Emerg Dis Published Online First: 2020. doi:10.1111/tbed.13684

55 Wehrhahn MC, Robson J, Brown S, et al. Self-collection: An appropriate alternative during the SARS-CoV-2 pandemic. J Clin Virol 2020;128:104417. doi:10.1016/j.jcv.2020.104417

56 Abdalhamid B, Bilder CR, McCutchen EL, et al. Assessment of Specimen Pooling to Conserve SARS CoV-2 Testing Resources. Am J Clin Pathol 2020;153:715–8. doi:10.1101/2020.04.03.20050195

57 Rao SN, Manissero D, Steele VR, et al. A Narrative Systematic Review of the Clinical Utility of Cycle Threshold Values in the Context of COVID-19. Infect Dis Ther 2020;:1–14. doi:10.1007/s40121-020-00324-3

58 Weemaes M, Martens S, Cuypers L, et al. Laboratory information system requirements to manage the COVID-19 pandemic: a report from the Belgian national reference testing center. J Am Med Inform Assoc Published Online First: April 2020. doi:10.1093/jamia/ocaa081

59 Liu Y, Liao W, Wan L, et al. Correlation Between Relative Nasopharyngeal Virus RNA Load and Lymphocyte Count Disease Severity in Patients with COVID-19. Viral Immunol Published Online First: 10 April 2020. doi:10.1089/vim.2020.0062

60 Brinati D, Campagner A, Ferrari D, et al. Detection of COVID-19 Infection from Routine Blood Exams with Machine Learning: A Feasibility Study. J Med Syst 2020;44:135. doi:10.1007/s10916-020-01597-4

61 Hamiel U, Kozer E, Youngster I. SARS-CoV-2 Rates in BCG-Vaccinated and Unvaccinated Young Adults. JAMA 2020;323:2340–1. doi:10.1001/jama.2020.8189

62 Chaisemartin C, Chaisemartin L. BCG vaccination in infancy does not protect against COVID-19. Evidence from a Natural Experiment. Clin Infect Dis Published Online First: 2020. doi:https://doi.org/10.1093/cid/ciaa1223

63 Van Damme W, Dahake R, Delamou A, et al. The COVID-19 pandemic: diverse contexts; different epidemics-how and why? BMJ Glob Heal 2020;5. doi:10.1136/bmjgh-2020-003098

